# Development and utilization of a surrogate SARS-CoV-2 viral neutralization assay to assess mRNA vaccine responses

**DOI:** 10.1101/2021.08.05.21261616

**Authors:** Adam V Wisnewski, Jian Liu, Carolina Lucas, Jon Klein, Akiko Iwasaki, Linda Cantley, Louis Fazen, Julian Campillo Luna, Martin Slade, Carrie A Redlich

## Abstract

**Background:** Tests for SARS-CoV-2 immunity are needed to help assess responses to vaccination, which can be heterogeneous and may wane over time. The plaque reduction neutralization test (PRNT) is considered the gold standard for measuring serum neutralizing antibodies but requires high level biosafety, live viral cultures and days to complete. We hypothesized that competitive enzyme linked immunoassays (ELISAs) based on SARS-CoV-2 spike protein’s receptor binding domain (RBD) attachment to its host receptor, the angiotensin converting enzyme 2 receptor (ACE2r), would correlate with PRNT, given the central role of RBD-ACE2r interactions in infection and published studies to date, and enable evaluation of vaccine responses.

**Methods and Findings:** Configuration and development of a competitive ELISA with plate-bound RBD and soluble biotinylated ACE2r was accomplished using pairs of pre/post vaccine serum. When the competitive ELISA was used to evaluate N=32 samples from COVID-19 patients previously tested by PRNT, excellent correlation in IC_50_ results were observed (r_s_= .83, *p* < 0.0001). When the competitive ELISA was used to evaluate N=41 vaccinated individuals and an additional N=14 unvaccinated recovered COVID-19 patients, significant differences in RBD-ACE2r inhibitory activity were associated with prior history of COVID-19 and type of vaccine received. In longitudinal analyses pre and up to 200 days post vaccine, surrogate neutralizing activity increased markedly after primary and booster vaccine doses, but fell substantially, up to <12% maximal levels within 6 months.

**Conclusions:** A competitive ELISA based on inhibition of RBD-ACE2r attachment correlates well with PRNT, quantifies significantly higher activity among vaccine recipients with prior COVID (vs. those without), and highlights marked declines in surrogate neutralizing activity over a 6 month period post vaccination. The findings raise concern about the duration of vaccine responses and potential need for booster shots.

## Introduction

Tests to evaluate SARS-CoV-2 immunity are need as vaccine responses and/or protection from prior infection can wane over time [1]. One surrogate of SARS-CoV-2 immunity is serum viral neutralizing antibodies, which are easy to access (venipuncture) and purify (centrifugation) and are stable if kept sterile and frozen [2–4]. Traditional tests for quantitating SARS-CoV-2 neutralizing antibodies (plaque reduction neutralization test or PRNT) are however, laborious, require biosafety level 3 (BSL3) working conditions due to use of live virus, and take several days to obtain results [5]. Rapid, less laborious tests for evaluating SARS-CoV-2 immunity will be especially helpful in better understanding the protection afforded by SARS-CoV-2 vaccination, including risk factors for suboptimal responses, and duration of the response.

The majority of neutralizing antibodies during natural SARS-CoV-2 infection target the receptor binding domain (RBD) of virus spike protein [6], which attaches to angiotensin converting enzyme 2 receptor (ACE2r) on airway epithelial cells to enter the host [7]. A number of monoclonal human antibodies that bind the SARS-CoV-2 spike protein RBD, but not all, are capable of inhibiting SARS-CoV-2 infection in vitro [7, 8]. Two RBD-specific human monoclonal antibodies have been developed for passive immunization therapy and shown positive therapeutic effects when administered early in the course of COVID-19 [9, 10].

Given the capacity of RBD-specific antibodies to block a critical primary step of infection (viral attachment to host ACE2r) [7] and their clinical efficacy in treatment [10, 11], in vitro assays that measure this activity may serve as surrogates of viral immunity and provide a measure of effective vaccine responsiveness. Competitive ELISAs that measure serum antibodies capable of blocking RBD-ACE2r binding have recently been described using samples from COVID-19 patients and correlate with live virus neutralization [12–15]. However, studies of surrogate neutralization assays in assessing vaccine responses have been limited [16].

The present investigation evaluated different competitive ELISA formats for quantifying human serum levels of antibodies that block ACE2r-RBD binding. Assay development with commercially available reagents was guided by empirical data using paired pre/post vaccine serum samples. The results of the optimized competitive ELISA were compared with previously published plaque reduction neutralization test (PRNT) results using live viral SARS-CoV-2, on N=32 samples from non-hospitalized, mild/asymptomatic COVID-19 patients [17]. The ACE2r-RBD competitive ELISA was then used to measure surrogate neutralizing activity post vaccination in N=41 individuals with distinct clinical profiles (history of COVID-19, vaccine type) compared to N=14 recovered COVID-19 patients subjects without vaccination. Changes in surrogate neutralizing activity over time up to 6 months post COVID-19 mRNA vaccination were also measured in serial samples from a limited number (N=4) of subjects. The results are discussed along with recently published data and concerns that mRNA vaccine efficacy may be waning and require a booster (third) dose [1, 18, 19].

## Materials and methods

### Human Subjects

The study used de-identified serum samples from prior studies [17, 20, 21] and new samples obtained from ongoing clinical studies. Subjects provided demographic information (age, race, sex) and 3cc of blood by venipuncture using vacutainer tubes (Becton Dickinson; Franklin Lake, NJ); serum was separated and stored frozen at −80°C until tested by enzyme linked immunosorbent assay (ELISA). The studies were reviewed, and ethical approval was given by the Yale University Institutional Review Board (IRB), protocols # 2000027810, 2000029735, and 2000027806. The Yale University IRB waived requirement for obtaining written consent from the participants given the research presented no more than minimal risk of harm and involved no procedures for which written consent is normally required outside of the research context. All participants provided informed verbal consent. The investigators explained the study to the subjects verbally, providing all pertinent information (purpose, procedures, risks, benefits, alternatives to participation, etc.), and allowed ample opportunity to ask questions. Following verbal explanation, the subject was provided with a study information sheet (written summary and secure web link) and afforded sufficient time to consider whether or not to participate in the research. After allowing the subject time to read the study information sheet, the investigators answered any additional questions before obtaining verbal agreement to participate in the research. The participants verbal agreement was noted and the time of entry into the study.

### Competitive ELISA

Ninety six well Nunc Maxisorp flat-bottom plates were coated with 50 μL/well of murine gamma-immunoglobulin constant region (mFc) dimerized SARS-CoV-2 (2019-nCoV) Spike Protein RBD (mFc-RBD) from Sino Biological (Chesterbrook, PA), catalog number: 40592-V05H, at 2 μg/mL in 1 x phosphate buffered saline (PBS) at 4°C overnight. Plates were blocked with 200 μL of 3% nonfat milk-PBS for 60 min at room temperature. Initial studies were performed with 2-100 ng/mL of human ACE2r labeled with biotin and detected with streptavidin-HRP (described below) to determine the linear range of the assay and an optimal concentration for inhibition assays. Subsequent competitive ELISAs were performed by pre-incubating wells with positive control, negative control, and experimental serum samples diluted from 1:10 to 1:800 in sample dilution buffer, which consisted of 0.05% Tween 20 in 1% milk-PBS. 50 µL each of the diluted controls and serum samples were added/well and incubated at room temperature for 30 minutes on a shaker. Subsequently, 50 µL of a human ACE2r labeled with biotin was added (40 ng/mL based on linear range in titration studies) and incubated at room temperature for 60 minutes on a shaker. After 5 washed with 0.05% Tween 20-PBS, 75 μL of streptavidin-HRP from ThermoFisher (Waltham, MA), catalog number N100 was added to each well and incubated for 60 minutes at room temperature. After 5 final washes, 100 μL of TMB substrate reagent from BD Biosciences (Franklin Lakes, NJ), catalog number #555214 was added to each well for ~15 minutes at room temperature, and reactions were terminated by addition of 50 μL of 2N HCl to each well. Plates were read at 450/655 nm. For quantitation and inter-assay standardization, all plates were run with an internal standard curve of ACE2r at varying concentrations ranging from 1.5 – 100 ng/mL, from which % inhibition and IC_50_ values were calculated. Based on data with control (pre-COVID and pre-vaccine) samples, IC_50_ values <1:10 were considered negative, and assigned an IC_50_ value of 1 for statistical calculations requiring log transformation described below.

### PRNT

Methods and data on PRNT of aliquots from the same samples used in this study have been previously published [17, 20].

### Statistics and software

All graphs were generated, and statistical analysis performed using Graph Pad Prism v 9.1.2 (GraphPad Software; San Diego, CA). Spearman rank order correlation was used to determine the association of IC_50s_ from PRNT and competitive ELISA data on paired samples. Significance differences in competitive ELISA data were calculated using the non-parametric Kruskal-Wallis tests and by ANOVA following log transformation of log-normal distributed IC_50_ values, with Tukey correction for multiple comparisons.

## Results

### Technical problems with competitive ELISA using soluble RBD and plate bound ACE2r

Initially we evaluated competitive ELISAs with plate bound ACE2r and soluble RBD to mimic in vivo biophysics and encountered technical difficulties consistent with cross-linkage of RBD in the soluble phase (see supplemental materials Fig S1 and S2). We hypothesize that RBD cross-linkage could be caused by antibodies that bind RBD but do not block ACE2r interaction [22], similar to certain human monoclonal antibodies [8], and consistent with the recent findings that plasmablast response to SARS-CoV-2 mRNA vaccination are dominated by non-neutralizing antibodies that target the RBD [23].

### Competitive ELISA with plate bound RBD and soluble ACE2r

Given technical issues possibly due to RBD cross-linking in competitive (ACE2r binding) ELISAs with serum from vaccinated individuals, we designed the reverse competitive ELISA, with plate bound RBD and soluble ACE2r (Fig 1). Competitive ELISAs were developed using murine IgG Fc domain tagged RBD and soluble ACE2r ectodomain tagged with biotin, with detection using streptavidin-labeled peroxidase. The results demonstrate dose dependent binding of ACE2r to plate bound RBD with an IC_50_ ~40 ng/mL and a wide linear range (Fig. 2).

**Figure 1.**
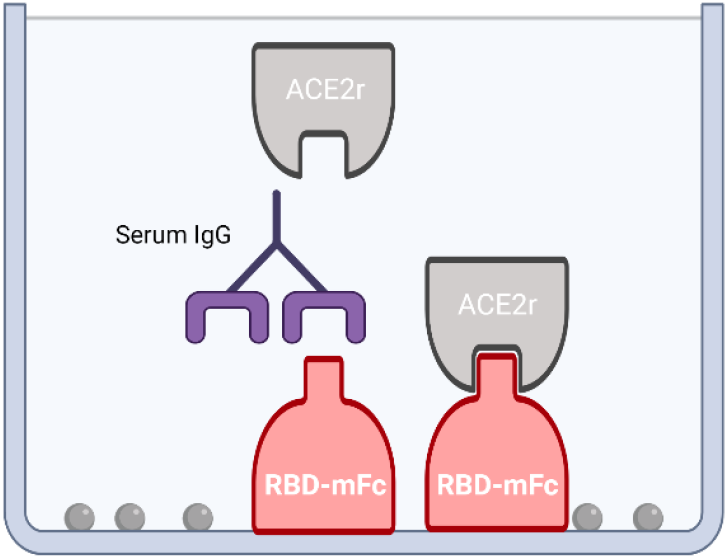
Competitive ELISA to detect antibodies that block SARS-CoV-2 spike-RBD binding to human ACE2r. A surrogate for neutralizing capacity is depicted with plate bound RBD-murine IgG Fc and soluble ACE2r.

**Figure 2.**
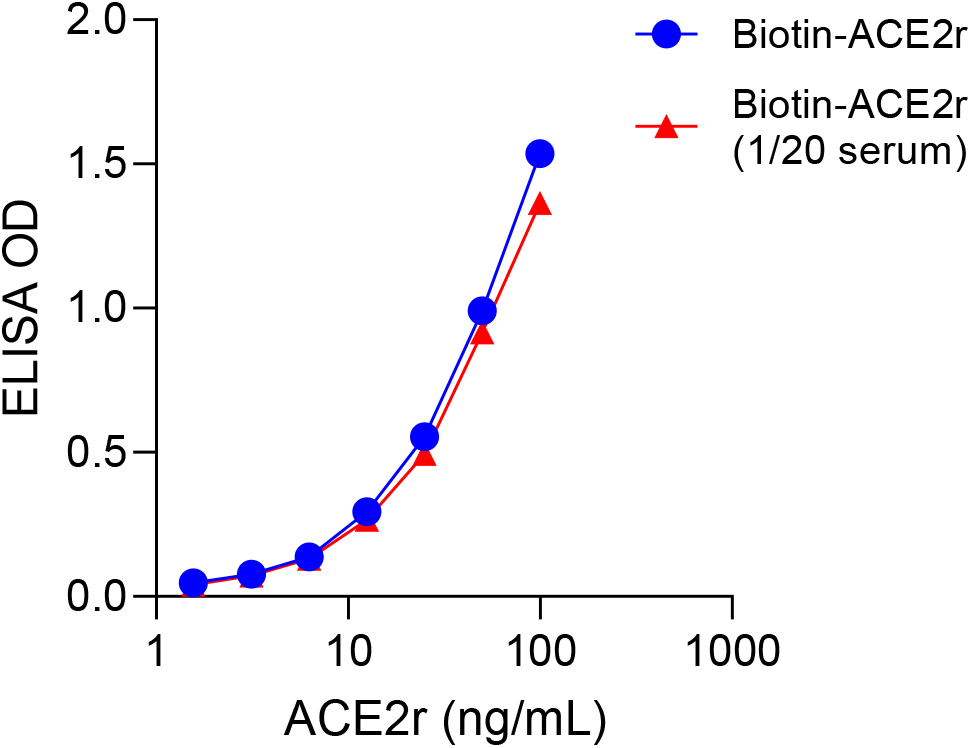
Soluble human ACE2r binding to plate bound SARS-CoV-2 antigen receptor binding domain (RBD). Biotin labeled human ACE2r (varying concentration in ng/mL on X-axis) was diluted in buffer or 1/20 serum (see key) and binding to RBD was quantified by ELISA (OD Y-axis) using HRP-labeled streptavidin.

When tested in the competitive ELISA, the inhibitory capacity of serum from N=3 individuals at a time of their optimal response post vaccination [21] was significantly higher than paired pre-vaccine serum samples, over a range of dilutions (Fig 3). All pre-vaccine samples demonstrated <50% inhibitory capacity at the highest serum concentration tested (1:10), suggesting an IC_50_ value <1:10 should be considered non-specific or negative.

**Figure 3.**
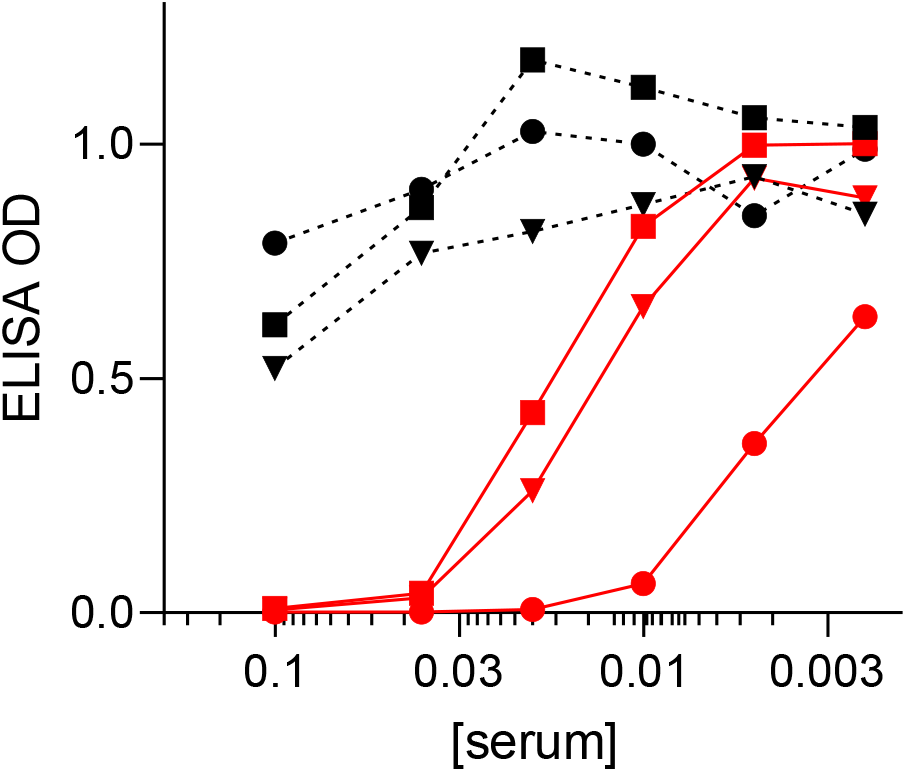
Surrogate SARS-COV-2 neutralization assays. Competitive RBD-ACE2r ELISAs with paired pre/post COVID-19 vaccine serum at different concentrations (X-axis) from different 3 subjects without prior COVID-19 before vaccine (black dashed lines) or day 7-10 post full vaccination (red solid lines). ELISA OD values reflecting RBD-ACE2r binding are shown on Y-axis. Subject 1 is square, subject 2 is triangle, and subject 3 is circle symbol.

### Competitive ELISA vs. PRNT

We next evaluated the competitive RBD-ACE2r ELISA as a surrogate neutralizing assay with N=32 serum samples from non-hospitalized COVID-19 patients. The samples demonstrated varying capacity to block biotinylated ACE2r binding to plate-bound RBD across a range of concentrations, from which IC_50_ values were calculated and compared with those obtained by PRNT using live SARS-CoV-2 [17, 20]. As shown in Fig 4, the IC_50s_ for serum inhibition of RBD-ACE2r binding showed highly significant (p < 0.0001) correlation (r_s_= 0.83) with corresponding IC_50_ values from titrated PRNT studies.

**Figure 4.**
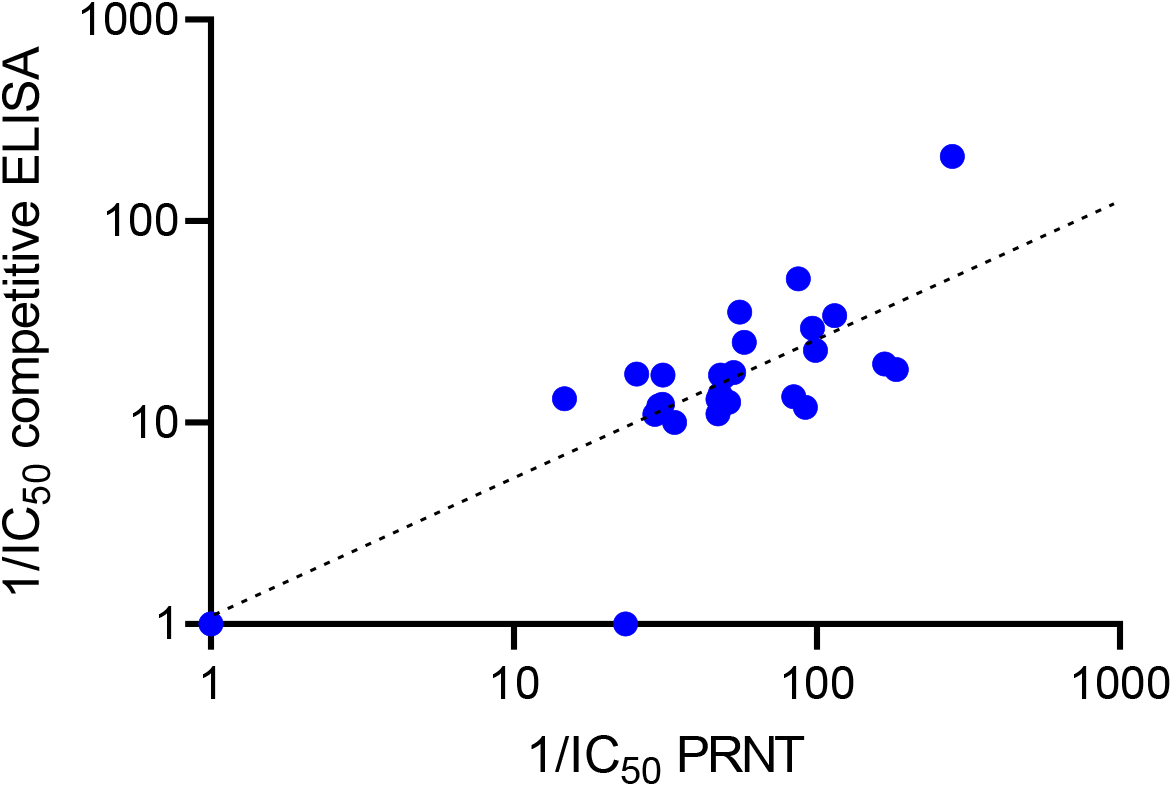
Correlation between PRNT and competitive ELISA. Each symbol represents the reciprocal IC_50_s from paired samples from the same subject tested in competitive ELISAs (Y-axis) and PRNT (X-axis). Note axes and trend line fitted to log scale.

### Surrogate neutralizing antibodies after infection vs. vaccination or both

We subsequently tested the neutralizing capacity of serum from N=54 individuals with 4 distinct profiles of infection and vaccination as described in Table 1. The subjects included COVID-19 patients without vaccination, COVID-19 subjects with vaccination, individuals without prior COVID-19, vaccinated with either Moderna or Pfizer-BioNTech mRNA vaccines, and paired baseline samples before COVID infection and or vaccination available for some subjects. The data demonstrate significant differences in IC_50_ values of serum samples depending on type of vaccine received and prior history of COVID-19 infection (Fig 5). The highest IC_50_ values were found among vaccinated individuals with prior COVID-19, with mean values significantly higher than those who received Pfizer-BioNTech (or had COVID-19 but no vaccine). Vaccinated subjects (Moderna and Pfizer-BioNTech) had significantly higher mean IC_50_ values than those with prior COVID-19, but no vaccination. Differences in RBD-ACE2r inhibitory activity were observed across a range of serum dilutions, suggesting a single serum dilution (rather than titration) may be sufficient for assessing vaccine responses (supplemental materials Fig S3). The observed heterogeneity in serum surrogate neutralizing activity among subjects within groups is likely due to multiple variables such as age or underlying medical conditions [24].

**Table 1.** Description of subjects with and without COVID-19 and/or mRNA vaccine.

| Group | Age (years)<br>(mean ± SD) | N | Vaccine/COVID-19<br>Status | Days after<br>COVID-19<br>(mean ± SD) | Days after<br>Vaccine<br>(mean ± SD) | Mean*<br>$IC_{50}$<br>(95% CI) |
| --- | --- | --- | --- | --- | --- | --- |
| A | 55 ± 11 | 16 | Pre-Vaccine / Pre-COVID-19 | - | - | 1 |
| B | 43 ± 11 | 13 | COVID-19 | 132 ± 43 | - | 12<br>(2-21) |
| C | 65 ± 11 | 14 | Pfizer-BioNTech | - | 45 ± 26 | 56<br>(27-87) |
| D | 64 ± 7 | 14 | Moderna | - | 55 ± 34 | 112<br>(63-160) |
| E | 54 ± 12 | 14 | COVID-19 + Vaccine | 127 ± 63 | 30 ± 26 | 195<br>(65-325) |

\* $p < 0.02$ for all groups except Moderna vs. COVID-19 + vaccine, and Moderna vs. Pfizer

**Figure 5.**
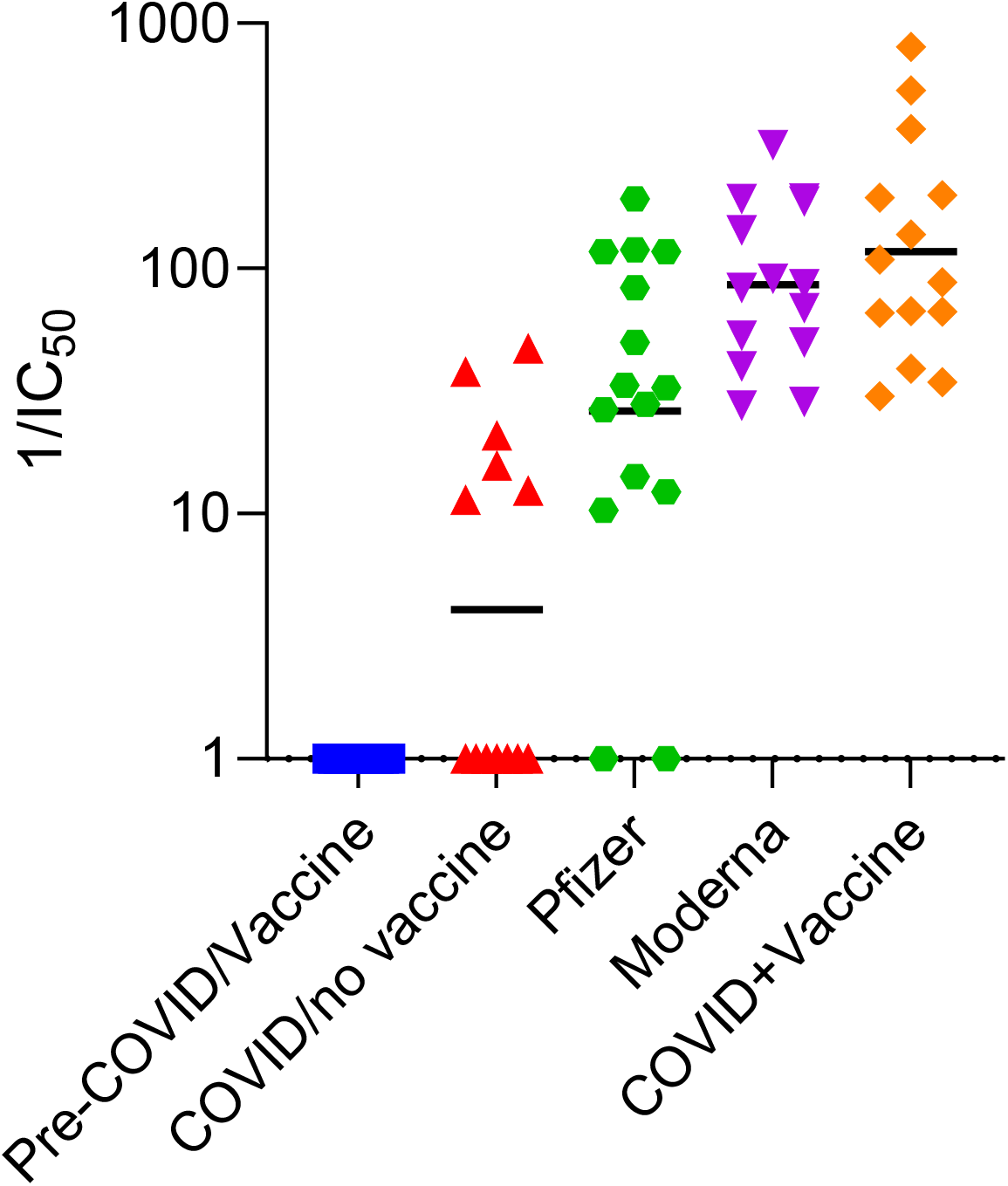
Inhibition of ACE2r binding to RBD by serum from different groups of individuals. The ability of different groups of individuals serum (described in Table 1) to inhibit ACE2r binding to RBD was measured by competitive ELISA and calculated reciprocal IC_50_ values are shown Y-axis. Data are from subject groups labeled in Table 1 as labeled. Each symbol represents a different person. Mean values are highlighted by bar in graph above and shown in Table 1, along with 95% CI and p values for groups with significant differences.

### Surrogate neutralizing antibodies over time post vaccination in individuals without COVID-19

We used the surrogate neutralization assay to evaluate serum samples collected pre-vaccine and serially over time up to 6 months after the 1^st^ dose of COVID-19 mRNA vaccine. Samples were from 4 subjects whose time course of serum IgA and IgG levels have recently been published [21]. As show in Fig 6, the IC_50_ for serum competition of RBD-ACE2r binding increased significantly after vaccination and remained near peak levels for approximately 50 days in all 4 subjects. However, a substantial proportion of neutralizing activity was lost by 200 days after (the 1^st^) vaccination, with <18% maximal levels remaining on average (range 11-27%).

**Figure 6.**
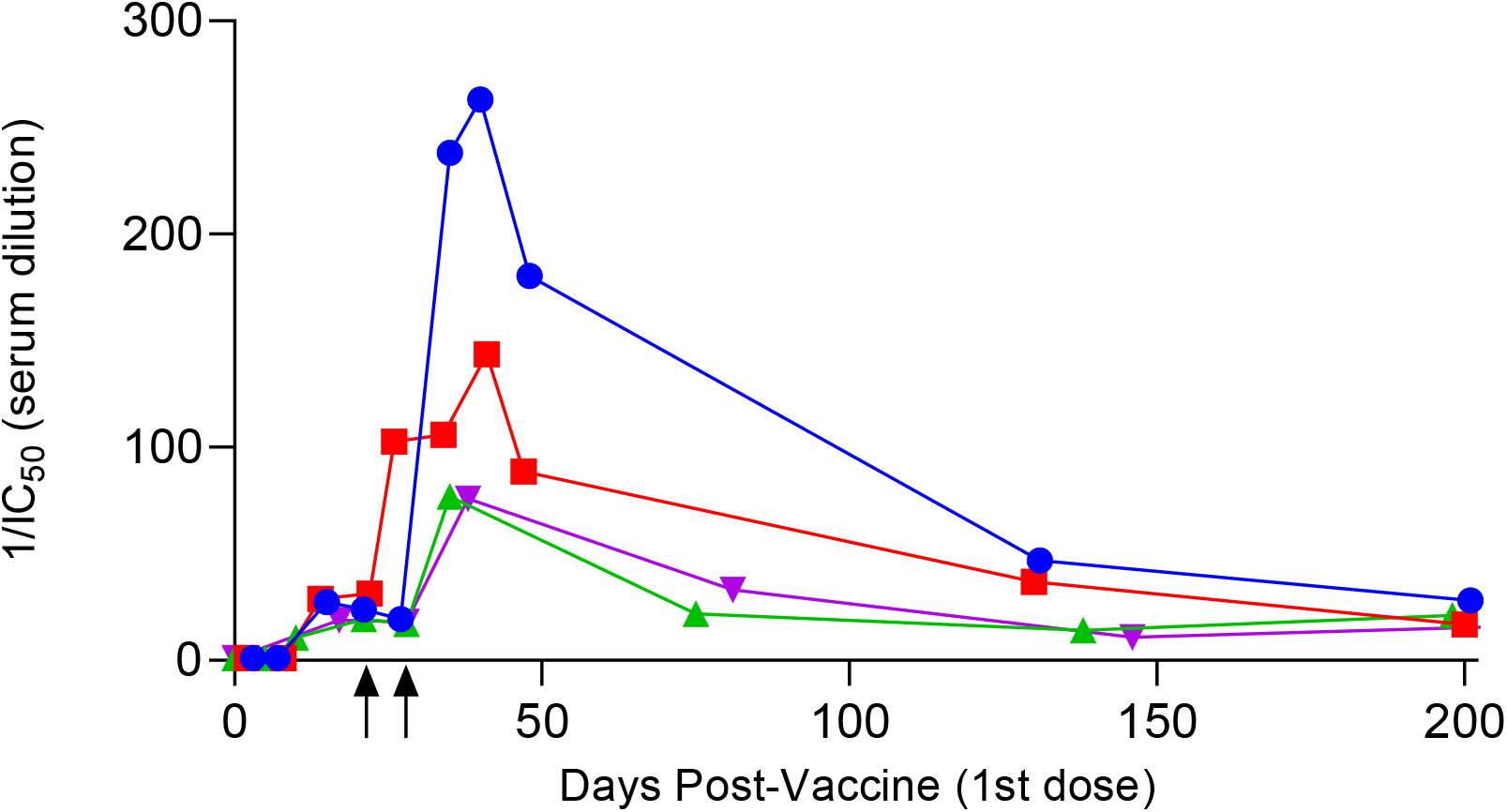
Changes in serum surrogate neutralizing activity over time after COVID-19 mRNA vaccination. The IC_50_ values for serial samples collected pre and up to 200 days post vaccination were calculated using the RBD-ACE2r competitive ELISA assay. Each symbol is a different individual previously described in Wisnewski et al Plos One 20201 [21]. Arrows on X-axis indicate timing of booster shots for subjects that received Pfizer-BioNTech (square), or Moderna (circle, triangle and inverted triangle). Note all samples were tested in duplicate and %CV < 10%.

## Discussion

The present study builds upon published data developing surrogate SARS-CoV-2 neutralization assays based on the binding of RBD to ACE2r, a critical step in the viral life cycle. We expanded upon this assays use assessing vaccine responses using well defined clinical samples, including available baseline samples from subjects prior to vaccination and/or COVID infection. We encountered technical issues in our initial assay design, which we hypothesize was due to vaccine-induction of antibodies that bind (cross-link) RBD but do not interfere with ACE2r interactions. To overcome these technical issues, we utilized the reverse formatted competitive ELISA (plate bound RBD-soluble ACE2r) to measure serum antibodies with neutralizing potential. The data demonstrate significant differences in surrogate neutralizing activity of individuals vaccinated after having had COVID-19 vs. unvaccinated subjects with prior COVID-19, and among subjects that received different vaccines (Pfizer-BioNTech vs. Moderna). The data also demonstrate a peak and slow decline in surrogate neutralizing antibodies from day 35 to 200 post initial vaccine dose.

The technical problems we encountered in competitive ELISAs with soluble RBD and plate bound ACE2r have not been reported to the best of our knowledge. However, some research use only kits for measuring RBD-ACE2r inhibiting antibodies are designed with plate bound RBD and soluble human ACE2r, without obvious explanation for this “reversed” orientation in regard to in vivo biophysics (e.g., cell surface bound ACE2r and soluble RBD). Recent work by Fenwick et al [25] in hospitalized COVID-19 patients using a soluble ACE2r-based binding format similar to the present study found strong correlations with PRNT, but did not study vaccinated individuals. The present data extend accumulating evidence that SARS-CoV-2 neutralizing capacity serum can be accomplished with competition assays based on soluble phase ACE2r attachment to immobilized spike or its RBD, and further demonstrate the utility of the approach in monitoring vaccine responses.

The present studies were performed with RBD encoded by the Wuhan-Hu-1 strain upon which the current mRNA vaccines are based, however RBDs and whole spike protein for the alpha, beta, gamma, and delta variants are currently available commercially and could be readily incorporated into the current competitive ELISA. A range of serum dilutions were tested in order to understand the dynamic range of the assay and to calculate IC_50_ values comparable to PRNT results. However, future screening of vaccine responses among large numbers of samples, might be effectively accomplished by measuring inhibition at a single (1:25) dilution, which we found most strongly correlation (r_s_ .76, *p* < 0.0001) with IC_50_ calculations.

The heterogeneity of surrogate neutralizing antibody responses among different subjects in the present study and the decline over time raise several concerns that may be relevant to the increasing numbers of vaccine breakthroughs, which have recently been reported and correlated with neutralizing antibody titers [1]. The current findings that vaccinated individuals with prior COVID-19 have higher levels of surrogate neutralizing activity than those without prior infection (or those infected without additional vaccination) imply that booster shots (e.g., a 3^rd^ exposure) may provide additional protection. A third exposure (e.g., 2^nd^ booster shot) is not uncommon for other vaccines to important human viruses such as polio, rotavirus, measles, mumps, rubella or bacteria [26–28]. The present data demonstrating a decline in surrogate neutralizing activity over 6 months post-vaccination further raise concern that SARS-CoV-2 vaccine protection is waning. Use of the present assay may be a feasible approach to assess SARS-CoV-2 protection of larger populations and identifying risk factors for suboptimal response to vaccine or waning responses over time.

The strengths and weaknesses of this study should be recognized in considering the significance of the findings. The major strength is the assay development with well-defined clinical samples, pre/post vaccine and/or COVID-19 samples, paired data from PRNT, samples from subjects that received different vaccines (Moderna, Pfizer-BioNTech) with different COVID-19 histories, and serial samples up to 6 months post vaccination. The major weakness of the study is the limited number of subjects per clinical group, which limited statistical power. In addition, the study was not designed to detect a correlation between neutralization activity and protection against infection, or involve children, whose immune responses may differ from those of adults. Further studies are urgently needed to better understand the heterogeneity and duration of COVID-19 vaccine-induced serum neutralizing responses in larger populations and their relationship to clinically relevant outcomes of SARS-CoV-2 exposure and infection.

In summary, we developed surrogate neutralization assay that significantly (p < 0.0001) correlates (r_*s*_=0.83) with IC_50_ measurements using live viral PRNT. The assay is similar in principle to methodology recently reported by Fenwick et al [25], but can be run with standard microtiter plate technology (vs. fluorescent microbeads) using commercially available reagents, and may provide an economical approach to evaluating vaccine responses and assessing immunity among large populations. Our initial data using the assay document heterogenous levels of surrogate neutralizing activity among vaccinated subjects and marked declines over a 6 month period. Together the data raise concern about the duration of vaccine induced protection from SARS-CoV-2 and potential need for booster shots.

## Supporting information

see supplemental materials Fig S1 and S2

## Data Availability

All data are available in the manuscript and/or supplemental materials.

## Notes

### Competing Interest Statement

The authors have declared no competing interest.

### Funding Statement

National Institute for Occupational Safety and Health, 5T03OH008607-15, to Dr. Carrie A Redlich.

### Author Declarations

The studies were reviewed, and ethical approval was given by the Yale University Institutional Review Board (IRB), protocols # 2000027810, 2000029735, and 2000027806. The Yale University IRB waived requirement for obtaining written consent from the participants given the research presented no more than minimal risk of harm and involved no procedures for which written consent is normally required outside of the research context. All participants provided informed verbal consent.

