## Supplementary material for "Development and utilization of a surrogate SARS-CoV-2 viral neutralization assay to assess mRNA vaccine responses": see supplemental materials Fig S1 and S2

Figure S1. RBD-ACE2r Competitive ELISA with plate bound ACE2r.

Figure S2. Schematic depiction of potential RBD cross-linkage in competitive ELSA

Figure S3. Inhibition of ACE2r binding to RBD by serum (1:25 dilution) from different groups of individuals.

Table S1. Information on study subjects

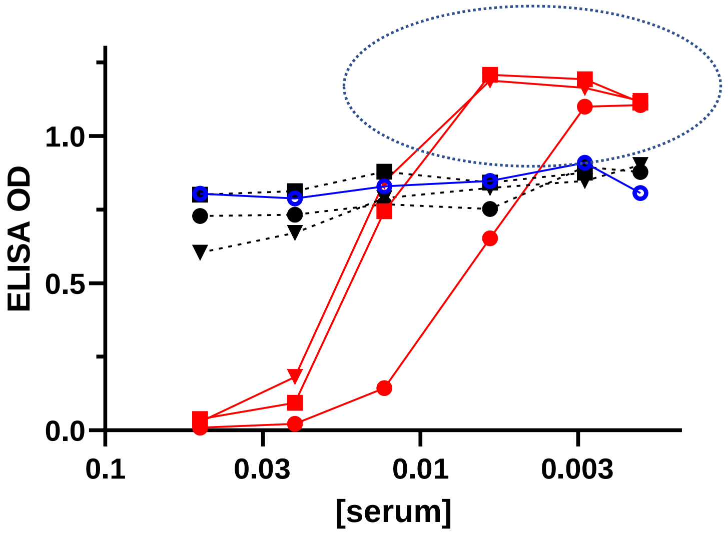


*Figure S1. RBD-ACE2r Competitive ELISA with plate bound ACE2r. RBD binding to ACE2r (Y-axis, reflected as optical density or OD) coated on microtiter plates in the presence of serum from pre (black dashed lines) or 7-10 days post 2nd dose of Moderna vaccine (red lines). Each symbol represents a different individual, subject 1 (circle), subject 2 (square) or subject 3 (triangle). Blue open circles is pooled serum from N=200 subjects pre-COVID era. Note increased binding at low serum concentrations in circled region, vs. control and pre-vaccine serum. The assay was designed with plate bound ACE2r and histidine labeled RBD, with detection by anti-histidine-HRP mAb, using ELISA methodology as described in the main text. Similar results were obtained with biotin labeled RBD and streptavidin-HRP detection*

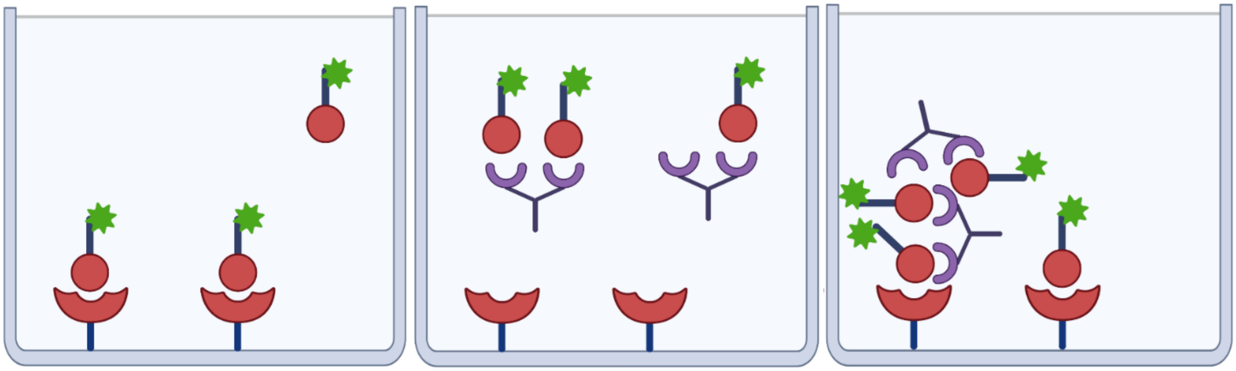

*Figure S2. Schematic depiction of potential RBD cross-linkage in competitive ELSA. Labeled RBD is depicted as *ball and stick and ACE2r is depicted bound to plate. Middle and right show antibody blocking binding or cross-linking RBD to ACE2r respectively.*

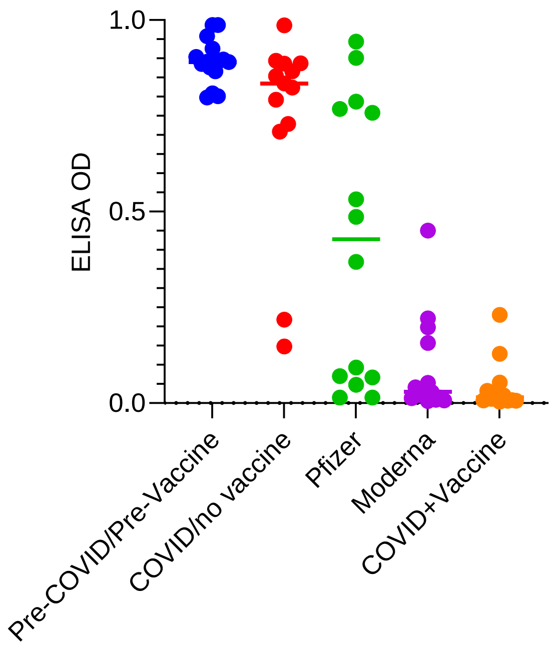


*Figure S3. Inhibition of ACE2r binding to RBD by serum (1:25 dilution) from different groups of individuals. The ability of different groups of individuals serum (described in Table 1 and Fig 5) to inhibit ACE2r binding to RBD was quantified. The ELISA OD (Y-axis), reflecting ACE2r binding was determined in the presence of a 1:25 dilution of serum. Each symbol represents a different subject.*

**S1 Table. Information on Study Subjects**

| | Patient ID | Sex | Age range | Infection Info | Vaccine Info | Days post 2nd dose | Spike OD | Surrogate Neut IC50 | PRNT IC50 |
|---|---|---|---|---|---|---|---|---|---|
| COVID (-), Vaccine-Moderna | | | | | | | | | |
| | 3024 | Male | 50-55 | COVID (-) | Moderna | 4 | 2.17 | 195 | |
| | 7082 | Female | 50-55 | COVID (-) | Moderna | 2 | 2.22 | 87 | |
| | 2737 | Male | 50-55 | COVID (-) | Moderna | 97 | 1.81 | 92 | |
| | 11001 | Male | 56-60 | COVID (-) | Moderna | 57 | 2.55 | 53 | |
| | 11002 | Female | 60-65 | COVID (-) | Moderna | 57 | 2.52 | 191 | |
| | 4602 | Male | 60-65 | COVID (-) | Moderna | 68 | 1.41 | 50 | |
| | 2446 | Female | 60-65 | COVID (-) | Moderna | 32 | 1.31 | 50 | |
| | 11024 | Male | 66-70 | COVID (-) | Moderna | 28 | 2.61 | 130 | |
| | 19010 | Male | 66-70 | COVID (-) | Moderna | 111 | 1.28 | 29 | |
| | 8114 | Female | 66-70 | COVID (-) | Moderna | 25 | 2.39 | 83 | |
| | 7211 | Female | 66-70 | COVID (-) | Moderna | 112 | 1.84 | 32 | |
| | 11008 | Male | 70-75 | COVID (-) | Moderna | 57 | 2.70 | 159 | |
| | 11004 | Female | 70-75 | COVID (-) | Moderna | 57 | 2.70 | 174 | |
| | 11025 | Female | 76-80 | COVID (-) | Moderna | 57 | 2.47 | 69 | |
| COVID (-), Vaccine-Pfizer | | | | | | | | | |
| | 2072 | Female | 56-60 | COVID (-) | Pfizer BioNTech | 2 | 0.41 | 11 | |
| | 19006 | Female | 30-35 | COVID (-) | Pfizer BioNTech | 126 | 1.39 | 33 | |
| | 1011 | Male | 46-50 | COVID (-) | Pfizer BioNTech | 18 | 1.18 | 90 | |
| | 9851 | Male | 50-55 | COVID (-) | Pfizer BioNTech | 77 | 1.00 | 36 | |
| | 9537 | Male | 50-55 | COVID (-) | Pfizer BioNTech | 54 | 0.49 | 1 | |
| | 1888 | Female | 56-60 | COVID (-) | Pfizer BioNTech | 61 | 1.98 | 194 | |
| | 1452 | Male | 56-60 | COVID (-) | Pfizer BioNTech | 55 | 1.67 | 28 | |
| | 1317 | Female | 56-60 | COVID (-) | Pfizer BioNTech | 69 | 1.35 | 50 | |
| | 7342 | Male | 66-70 | COVID (-) | Pfizer BioNTech | 76 | 1.08 | 1 | |
| | 17230 | Female | 66-70 | COVID (-) | Pfizer BioNTech | 10 | 1.63 | 88 | |
| | 7042 | Male | 70-75 | COVID (-) | Pfizer BioNTech | 67 | 2.18 | 84 | |
| | 1346 | Male | 70-75 | COVID (-) | Pfizer BioNTech | 10 | 2.35 | 117 | |
| | 19041 | Male | 70-75 | COVID (-) | Pfizer BioNTech | 63 | 1.84 | 14 | |
| | 12060 | Female | 80-85 | COVID (-) | Pfizer BioNTech | 27 | 1.36 | 27 | |
| | 5818 | Male | 80-85 | COVID (-) | Pfizer BioNTech | 37 | 1.04 | 12 | |
| COVID (+), Vaccine-Moderna | | | | | | | | | |
| | 2315 | Male | 36-40 | 73 days | Moderna | 57 | 1.93 | 39 | |
| | 17212 | Female | 40-45 | 108 days | Moderna | 21 | 2.24 | 67 | |
| | 6107 | Male | 46-50 | 123 days | Moderna | 17 | 2.41 | 193 | |
| | 2097 | Male | 50-55 | 119 days | Moderna | 7 | 1.84 | 34 | |
| | 3708 | Female | 50-55 | NA | Moderna | 22 | 2.48 | 67 | |
| | 17005 | Male | 50-55 | 215 days | Moderna | 37 | 2.58 | 88 | |
| | 8943 | Male | 56-60 | 234 days | Moderna | 99 | 2.24 | 66 | |
| | 11027 | Female | 65-70 | 90 days | Moderna | 7* | 2.85 | 800 | |
| COVID (+), Vaccine-Pfizer | | | | | | | | | |
| | 4318 | Male | 30-35 | 211 days | Pfizer BioNTech | 19 | 2.72 | 30 | |
| | 4778 | Male | 40-45 | 62 days | Pfizer BioNTech | 9 | 2.22 | 198 | |
| | 9378 | Female | 60-65 | 139 days | Pfizer BioNTech | 53 | 2.30 | 109 | |
| | 18056 | Female | 66-70 | 149 days | Pfizer BioNTech | 7 | 2.33 | 544 | |
| | 18079 | Female | 66-70 | 82 days | Pfizer BioNTech | 22 | 2.51 | 137 | |
| | 17228 | Male | 70-75 | 80 days | Pfizer BioNTech | 8 | 2.37 | 378 | |
| COVID (+), Vaccine (-) | | | | | | | | | |
| | 19049 | Male | 20-25 | 163 days | No vaccine | | 1.63 | 11 | |
| | 2546 | Male | 30-35 | 75 days | No vaccine | | 1.86 | 1 | |
| | 3010 | Female | 30-35 | 186 Days | No vaccine | | 1.09 | 10 | |
| | 4261 | Male | 36-40 | 51 days | No vaccine | | 0.77 | 1 | |
| | 17227 | Female | 40-45 | 156 days | No vaccine | | 1.45 | 1 | |
| | 1616 | Female | 40-45 | 70 Days | No vaccine | | 2.54 | 21 | |
| | 10088 | Male | 40-45 | 100 days | No vaccine | | 1.64 | 1 | |
| | 19051 | Male | 46-50 | 132 days | No vaccine | | 1.81 | 12 | |
| | 17107 | Male | 46-50 | 336 days | No vaccine | | 1.65 | 11 | |
| | 18098 | Female | 46-50 | 185 days | No vaccine | | 2.44 | 47 | |
| | 18036 | Male | 46-50 | 62 days | No vaccine | | 1.18 | 1 | |
| | 9926 | Female | 50-55 | 174 days | No vaccine | | 1.17 | 1 | |
| | 1606 | Male | 60-65 | 24 days | No vaccine | | 3.00 | 38 | |
| COVID (-), Vaccine (-) | | | | | | | | | |
| | 3010 | Female | 30-35 | COVID (-) | pre Vaccine | | 0.04 | 1 | |
| | 4261 | Male | 36-40 | COVID (-) | pre Vaccine | | 0.14 | 1 | |
| | 1616 | Female | 40-45 | COVID (-) | pre Vaccine | | 0.10 | 1 | |
| | 6753 | Female | 40-45 | COVID (-) | pre Vaccine | | 0.04 | 1 | |
| | 7082 | Female | 50-55 | COVID (-) | pre Vaccine | | 0.31 | 1 | |
| | 7082 | Female | 50-55 | COVID (-) | pre Vaccine | | 0.28 | 1 | |
| | 9851 | Male | 50-55 | COVID (-) | pre Vaccine | | 0.30 | 1 | |
| | 8943 | Male | 56-60 | COVID (-) | pre Vaccine | | 0.06 | 1 | |
| | 9704 | Female | 56-60 | COVID (-) | pre Vaccine | | 0.43 | 1 | |
| | 1317 | Female | 56-60 | COVID (-) | pre Vaccine | | 0.06 | 1 | |
| | 9378 | Female | 60-65 | COVID (-) | pre Vaccine | | 0.12 | 1 | |
| | 4602 | Male | 60-65 | COVID (-) | pre Vaccine | | 0.24 | 1 | |
| | 2446 | Female | 60-65 | COVID (-) | pre Vaccine | | 0.09 | 1 | |
| | 7342 | Male | 66-70 | COVID (-) | pre Vaccine | | 0.13 | 1 | |
| | 7211 | Female | 66-70 | COVID (-) | pre Vaccine | | 0.05 | 1 | |
| | 7042 | Male | 70-75 | COVID (-) | pre Vaccine | | 0.22 | 1 | |
| PRNT samples | N1 | Male | 20-25 | | No vaccine | | 0.783 | 13 | 31 |
| | N2 | Male | 20-25 | | No vaccine | | 1.154 | 1 | 48 |
| | N3 | Female | 20-25 | | No vaccine | | 0.813 | 18 | 49 |
| | N4 | Male | 20-25 | | No vaccine | | 0.9425 | 1 | 48 |

| | | | | | | |
|---|---|---|---|---|---|---|
| N5 | Male | 20-25 | No vaccine | 1.56 | 1 | 34 |
| N6 | Male | 20-25 | No vaccine | 1.344 | 20 | 30 |
| N7 | Female | 20-25 | No vaccine | 0.7435 | 17 | 25 |
| N8 | Male | 20-25 | No vaccine | 1.0185 | 14 | 183 |
| N9 | Male | 20-25 | No vaccine | 1.4225 | 25 | 168 |
| N10 | Male | 20-25 | No vaccine | 0.5015 | 12 | 64 |
| N11 | Male | 20-25 | No vaccine | 0.5205 | 35 | 47 |
| N12 | Male | 20-25 | No vaccine | 1.4385 | 34 | 97 |
| N13 | Male | 20-25 | No vaccine | 0.7555 | 1 | 92 |
| N14 | Male | 20-25 | No vaccine | 2.171 | 13 | 87 |
| N15 | Male | 20-25 | No vaccine | 0.8775 | 13 | 84 |
| N16 | Male | 20-25 | No vaccine | 1.963 | 11 | 56 |
| N17 | Male | 20-25 | No vaccine | 1.0905 | 12 | 15 |
| N18 | Female | 26-30 | No vaccine | 0.663 | 13 | 51 |
| N19 | Male | 26-30 | No vaccine | 0.516 | 1 | 1 |
| N20 | Male | 26-30 | No vaccine | 0.9935 | 12 | 29 |
| N21 | Male | 26-30 | No vaccine | 1.5045 | 18 | 179 |
| N22 | Male | 26-30 | No vaccine | 1.2155 | 52 | 58 |
| N23 | Male | 26-30 | No vaccine | 0.423 | 17 | 1 |
| N24 | Male | 30-35 | No vaccine | 1.264 | 1 | 99 |
| N25 | Male | 40-45 | No vaccine | 0.0535 | 17 | 1 |
| N26 | Male | 40-45 | No vaccine | 2.9905 | 10 | 281 |
| N27 | Female | 40-45 | No vaccine | 0.9525 | 17 | 115 |
| N28 | Male | 46-50 | No vaccine | 1.034 | 209 | 53 |
| N29 | Male | 50-55 | No vaccine | 0.77 | 29 | 47 |
| N30 | Male | 50-55 | No vaccine | 0.66 | 1 | 23 |
| N31 | NA | NA | No vaccine | 0.0815 | 23 | 1 |
| N32 | NA | NA | No vaccine | 0.0345 | 11 | 1 |

* = days after 1st dose
NA = not available
